# Procarbazine-induced Genomic Toxicity in Hodgkin Lymphoma Survivors

**DOI:** 10.1101/2024.06.04.24308149

**Authors:** Anna Santarsieri, Emily Mitchell, My H. Pham, Rashesh Sanghvi, Janina Jablonski, Henry Lee-Six, Katherine Sturgess, Pauline Brice, Tobias F. Menne, Wendy Osborne, Thomas Creasey, Kirit M. Ardeshna, Joanna Baxter, Sarah Behan, Kaljit Bhuller, Stephen Booth, Nikesh D. Chavda, Graham P. Collins, Dominic J. Culligan, Kate Cwynarski, Andrew Davies, Abigail Downing, David Dutton, Michelle Furtado, Eve Gallop-Evans, Andrew Hodson, David Hopkins, Hannah Hsu, Sunil Iyengar, Stephen G. Jones, Mamatha Karanth, Kim M. Linton, Oliver C. Lomas, Nicolas Martinez-Calle, Abhinav Mathur, Pamela McKay, Sateesh K. Nagumantry, Elizabeth H. Phillips, Neil Phillips, John F. Rudge, Nimish K. Shah, Gwyneth Stafford, Alex Sternberg, Rachel Trickey, Benjamin J. Uttenthal, Natasha Wetherall, Xiao-Yin Zhang, Andrew K. McMillan, Nicholas Coleman, Michael R. Stratton, Elisa Laurenti, Peter Borchmann, Sven Borchmann, Peter J. Campbell, Raheleh Rahbari, George A. Follows

## Abstract

**Background:** Procarbazine-containing chemotherapy regimens associate with cytopenias and infertility, suggesting stem cell toxicity. Procarbazine in eBEACOPP (escalated dose bleomycin, etoposide, doxorubicin, cyclophosphamide, vincristine, procarbazine, prednisolone) is increasingly replaced with dacarbazine (eBEACOPDac) to reduce toxicity, although limited genomic and clinical data support this substitution.

**Methods:** To assess mutagenic and clinical consequences of dacarbazine-procarbazine substitutions, we compared mutational landscapes in haematopoietic stem and progenitor cells (HSPCs) from patients treated with different Hodgkin regimens and children, sperm and bowel tissue from procarbazine-treated patients. We compared efficacy and toxicity data of a multicentre eBEACOPDac-treated patient cohort, with eBEACOPP clinical trial and real-world datasets.

**Results:** eBEACOPP-treated patients exhibit a higher burden of point mutations, small insertions and deletions in HSPCs compared to eBEACOPDac and ABVD (doxorubicin, bleomycin, vinblastine, dacarbazine)-treated patients. Two novel mutational signatures, SBSA (SBS25-like) and SBSB were identified in HSPCs, neoplastic and normal colon from only procarbazine-treated patients. SBSB was also identified in germline DNA of three children conceived post-eBEACOPP and sperm of an eBEACOPP-treated male. The dacarbazine substitution did not appear to compromise efficacy; 3-year progression-free survival of 312 eBEACOPDac patients (93.3%; CI_95_=90.3-96.4%) mirrored that of 1945 HD18-trial eBEACOPP patients (93.3%; CI_95_=92.1-94.4%). eBEACOPDac-treated patients required fewer blood transfusions, demonstrated higher post-chemotherapy sperm concentrations, and experienced earlier resumption of menstrual periods.

**Conclusions:** Procarbazine induces a higher mutational burden and novel mutational signatures in eBEACOPP-treated patients and their germline DNA raising concerns for hereditary consequences. However, replacing procarbazine with dacarbazine appears to mitigate gonadal and stem cell toxicity while maintaining comparable clinical efficacy.

---

Classical Hodgkin lymphoma (cHL) is highly curable with modern therapies. Escalated BEACOPP (bleomycin, etoposide, doxorubicin, cyclophosphamide, vincristine, procarbazine and prednisolone at escalated doses; eBEACOPP) is a gold standard first-line treatment for advanced-stage Hodgkin lymphoma conferring superior long-term progression-free survival compared with other polychemotherapy regimens^1–3^. However, there are significant short- and long-term side-effects, including gonadal and stem cell toxicity. Combination chemotherapy protocols containing procarbazine are known to cause dose-dependent infertility^4,5^, and procarbazine has been shown to have mutagenic effects in animal models^6^. Replacing procarbazine with dacarbazine in COPP reduced gonadal toxicity and conferred comparable long-term event-free survival in paediatric cHL^7^.

While the late effects of eBEACOPP have been well documented^8–10^, the mutational impact on stem cells is less well defined. Certain chemotherapy agents are known to induce specific mutational signatures in cancer cells. In the Catalogue of Somatic Mutations in Cancer (COSMIC) database^11^, chemotherapy is proposed to be responsible for 7 out of 67 mutational signatures, with the aetiology of many other mutational signatures yet to be defined. Chemotherapy-induced mutagenesis has also been reported in normal somatic cells, including colon stem cells^12^ and peripheral blood cells^13^. Mutational signatures caused by platinum and alkylating agents have been identified in the germline of children with paternal chemotherapy exposure prior to conception^14^.

To investigate the genomic impact of cHL therapies in normal stem cells, we compared the mutational burden and signatures in the haematopoietic stem and progenitor cells (HSPCs) of long-term remission patients previously treated with doxorubicin, bleomycin, vinblastine, dacarbazine (ABVD), eBEACOPP or escalated BEACOPDac (eBEACOPDac) where procarbazine was replaced by dacarbazine. We then defined the extent of chemotherapy-induced mutagenesis in other stem cell compartments and in tumour and normal tissues from one patient. To study the impact of eBEACOPP on both maternal and paternal gonadal stem cells, we mapped mutational signatures and burdens in 5 children of a female patient (two conceived pre- and three post-eBEACOPP) as well as in sperm from a male donor treated with eBEACOPP.

To validate the clinical efficacy of the eBEACOPDac regimen we collected multicentre data on eBEACOPDac-treated patients and compared clinical outcomes with two independent eBEACOPP datasets: the German HD18 trial to compare treatment efficacy and maximise statistical power, and a real-world UK eBEACOPP dataset to compare specific toxicity outcomes.

## Methods

### Mutational burden and signature analysis

Peripheral blood mononuclear cells (PBMC) were isolated from 12 advanced-stage Hodgkin lymphoma patients who had been in remission for ≥6 months. The patients had been previously treated with either eBEACOPDac (n=4), eBEACOPP (n=5) or ABVD (n=3). PBMCs were cultured for 14 days and single-cell derived HSPC colonies were harvested. DNA was extracted from each colony and 6-8 single-cell derived HSPC colonies were whole-genome sequenced from each of the 12 patients (n=91; mean sequencing depth 26X).

CaVEMan^15^, used for calling single nucleotide variants (SNVs), was run against an unmatched synthetic normal genome. Insertions and deletions (indels) were identified using the Pindel algorithm applied to a matched normal sample.

The linear regression of age and SNV or indel mutation burden^16^ from a control cohort (n=110; mean sequencing depth 24X) was used as a baseline against which to compare the mutation burden of the chemotherapy-exposed individuals.

Whole-genome sequencing (WGS) was performed on a caecal adenocarcinoma from a Hodgkin lymphoma survivor treated with Chlorambucil, Vinblastine, Procarbazine, Prednisolone (ChlVPP) nine years before sampling^17^. Previously published^17^ normal colorectal epithelium from the same individual was also interrogated.

Buccal DNA was obtained from five children and the spouse of a cHL female patient (who was treated with eBEACOPP prior to conceiving her 3rd child) and subjected to WGS (Supplementary Methods). Mutational burden and signatures were compared between pre-(n=2) and post-chemotherapy children (n=3).

The sperm DNA of a patient with mild oligospermia (13M/ml) 3.5 years post 4 cycles of eBEACOPP treatment was sequenced using Nanoseq WGS^18^ (Supplementary Methods). To establish a control cohort for comparison, the parental germline de novo point mutation burden (DNM) was predicted based on pedigree studies from parents and offspring trio WGS^19^.

The Hierarchical Dirichlet process (HDP) was employed to extract mutational signatures from SBS/Indels derived from HSPCs, germline DNMs from the offspring of an eBEACOPP-treated female, and mutations from the sperm of an eBEACOPP-treated male (Supplementary Methods).

### Clinical data analysis

This is a retrospective study of 312 patients with advanced-stage cHL, treated with first-line escalated BEACOPDac (Table S1) at 25 centres in the UK, Ireland and France between 2017 and 2022.

Progression-free survival (PFS) of the 312 eBEACOPDac-treated patients were compared with 1945 eBEACOPP-treated HD18 trial patients^20^, through collaboration with the German Hodgkin Study Group (GHSG) (Supplementary Methods).

Toxicity outcomes of the eBEACOPDac cohort were compared with a real-world eBEACOPP cohort of 73 patients treated at 7 UK centres between 2009 and 2022. The outcomes studied were day 8 (D8) neutrophil count, D8 alanine transaminase (ALT), units of red cells transfused, and days of non-elective hospital admission, time to return of menstrual periods in females aged <35 years, and sperm concentration pre- and ≥2 years post-chemotherapy.

Statistical analyses were performed using R software and SPSS. The Mann-Whitney U test and t-test were used for continuous variables and Fisher’s exact test for categorical variables. The study was conducted with Health Research Authority and Public Benefit and Privacy Panel approval.

## Results

### Mutational burden

We found a linear correlation between both the number of SNV and indel burdens with age across all cohorts (Figure 1A). In line with previous studies, normal adult HSPCs accumulated ∼18 SNVs per year post birth^16^. Consequently, the mutation burden increases from 400 SNVs in 20-year-old individuals to nearly 1500 SNVs at the age of 80 years. Additionally, HSPCs from all chemotherapy-exposed individuals demonstrated higher than expected mutation loads for their respective ages. ABVD-treated patients and eBEACOPDac-treated patients had a minor excess somatic mutation burden of 186 (CI_95_=116-254) and 290 (CI_95_=241-339), respectively, compared to age-matched normal HSPCs. However, we observed significant elevations in SNV burdens in patients receiving eBEACOPP treatment, with 1150 (CI_95_=934-1366) excess mutations. A similar pattern was also observed while analysing small indels in HSPCs. There was a comparative indel mutation burden of −0.4 (CI_95_=-2.9-2.1) in ABVD-treated patients, 11.2 (CI_95_=6.9-15.6) in eBEACOPDac-treated patients and 64.8 (CI_95_=53.1-76.5) in eBEACOPP-treated patients (Extended Figure 1A).

**Figure 1:**
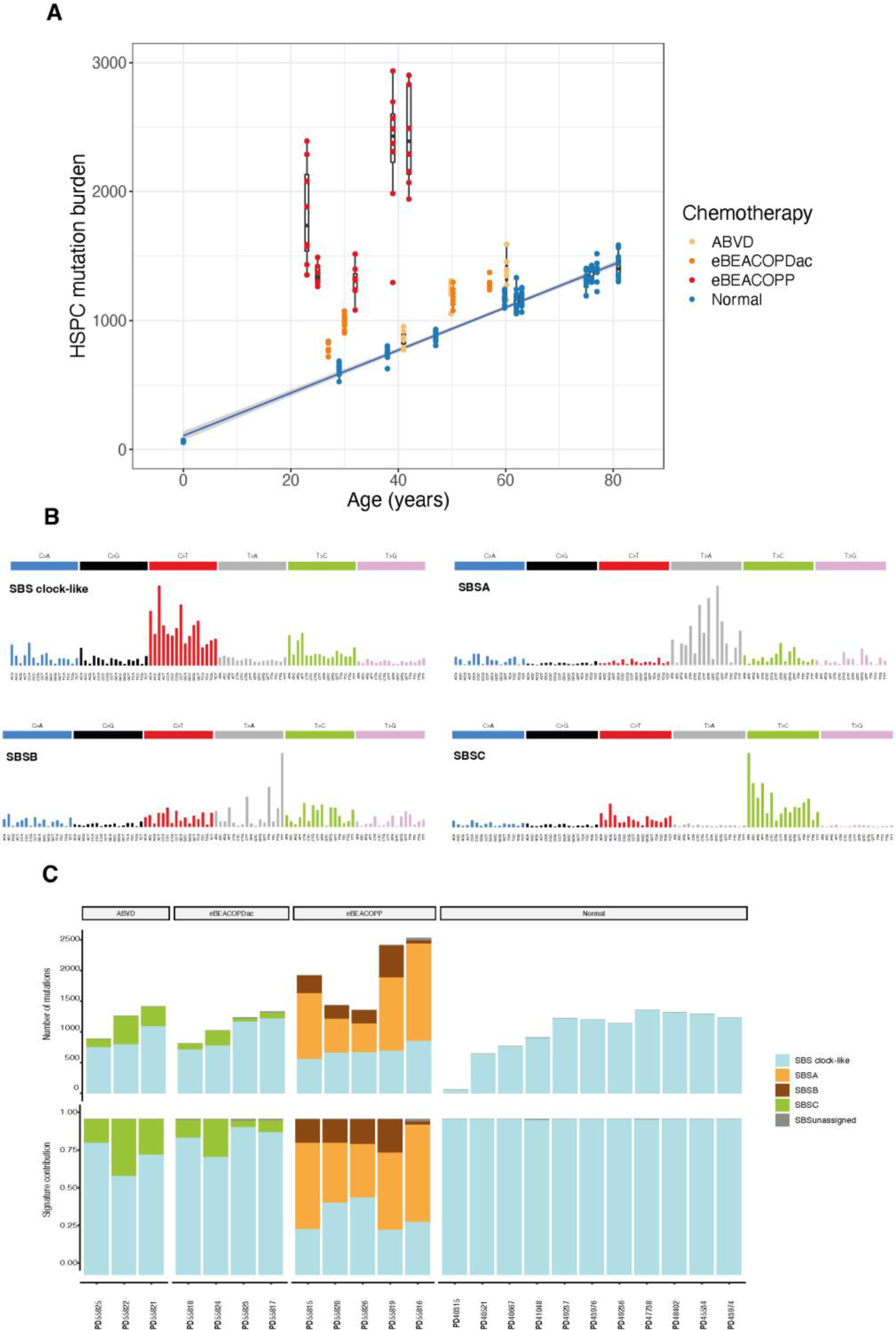
**(A)** Burden of HSPC single nucleotide variants across the chemotherapy donor and a comparable normal donor cohort. The boxplots represent data from individual HSPC colonies (n = 201; 6-10 colonies per individual) and are coloured by chemotherapy exposure. The boxes indicate the median and interquartile range and the whiskers denote the range. The blue line represents a regression of age on mutation burden for the normal donors, with 95% CI shaded in grey. **(B)** Mutational signatures extracted using HDP. **(C)** Proportion of extracted signatures active in eBEACOPP, cBEACOPDac, ABVD and normal individuals. HSPC = Haematopoietic stem and progenitor cells, HDP= Hierarchical Dirichlet process

The post-chemotherapy children had a modest but significant number of de novo germline point mutations (DNMs). After correcting for the paternal component of DNMs, we observed a significant increase (∼2.3 fold; Supplementary Figure 3) in the maternally inherited DNMs among the post-chemotherapy children (p-value 0.02505; t-test; Figure 2C). Moreover, analysis of sperm DNA of a patient post-eBEACOPP revealed a 3-fold higher mutation burden, after correcting for his age, compared to the control cohort^19^ (Methods; Figure 2D).

**Figure 2:**
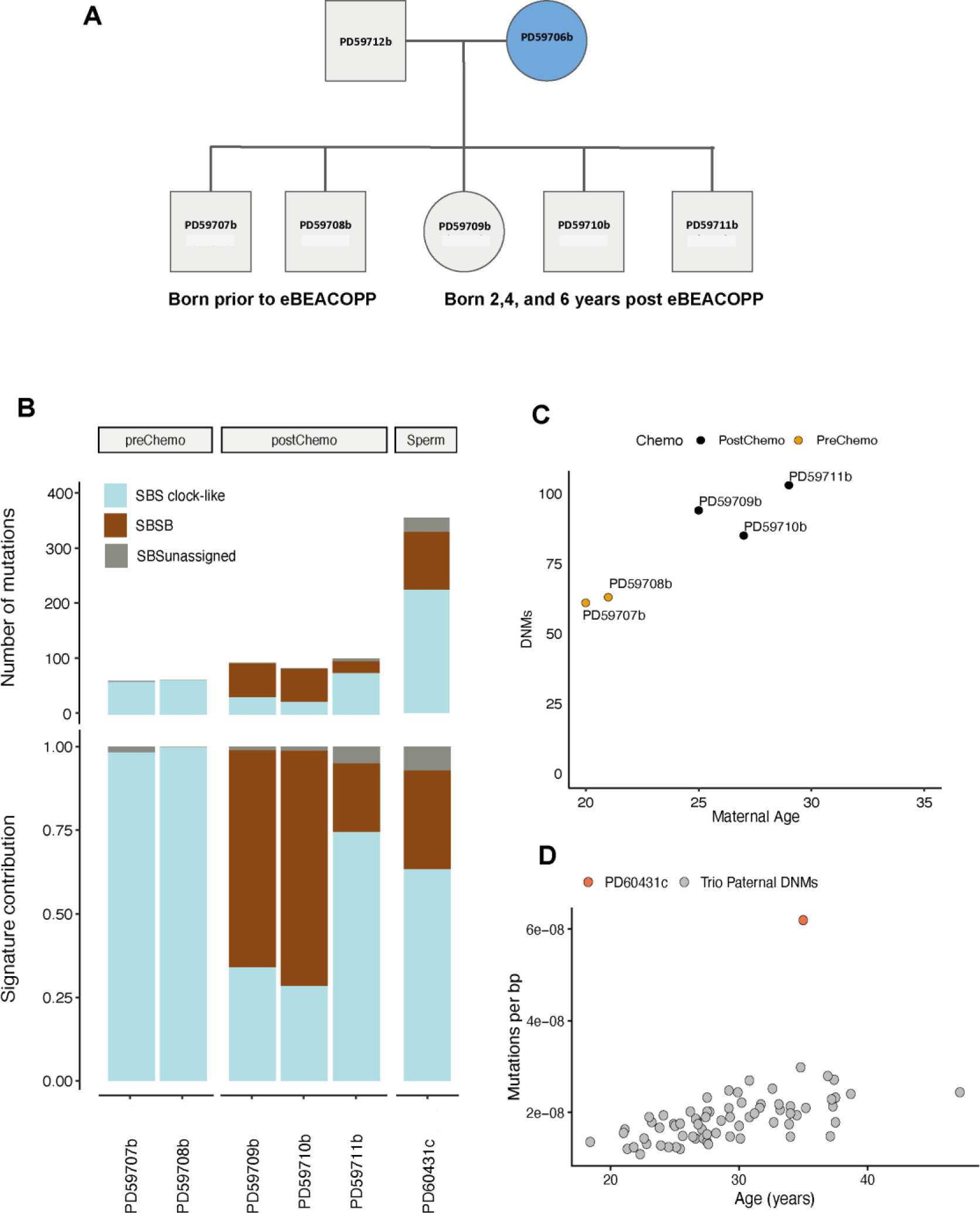
**(A)** Pedigree of female Hodgkin lymphoma patient who received 6 cycles eBEACOPP. 2 children were born pre-chemotherapy, and 3 children were conceived post-chemotherapy. **(B)** SBS mutational signature contribution in pre- and post-chemotherapy children and sperm sample from the patient treated with 4 cycles eBEACOPP. **(C)** Number of *de novo* mutations in pre- and post-chemotherapy children plotted against maternal age (p-value 0.01109). P-value was generated using t-test to check significance of the pre-chemo probands burden to the post-chemo probands. **(D)** Mutation burden in sperm of oligospermic patient treated with 4 cycles eBEACOPP compared with paternal mutation burden in trio studies^10^. DNMs = De novo mutations. SBS = single base substitution. Bp = base pair

### Mutational Signature analysis

Four SNV mutational signatures were extracted from HSPCs, namely: SBS clock-like, SBSA, SBSB, and SBSC (Methods; Figure 1C). Other than SBS clock-like signature, the remaining three signatures are novel and have not been reported in the COSMIC database. SBS clock-like is a combination of SBS1, associated with the deamination of 5-methylcytosine, SBS5 of unknown aetiology, and ‘SBSBlood’, the predominant ‘clock-like’ signature present in HSPCs. SBSA is only observed in eBEACOPP treated patients and shares 0.84 cosine similarity to COSMIC v3.4 signature SBS25. Similarly, SBSB, also with predominant T>A substitutions, is found in all patients exposed to eBEACOPP, but with smaller contributions. These two mutational signatures have been recently ascribed to procarbazine treatment^21,22^. SBSC was detected in patients treated with either ABVD or eBEACOPDac (Figure 1C). The sole shared chemotherapy agent in these protocols is Dacarbazine, implying an association between this drug and SBSC. Specific SBS mutational signatures were not obviously associated with topoisomerase inhibitors, vinca-alkaloids or bleomycin.

Two indel signatures were extracted, of which one signature was a combination of the COSMIC v3.4 indel clock-like signatures. The second signature was a novel signature that was observed only in eBEACOPP-treated patients and recently attributed to procarbazine^21^ (Extended Figure 1B, 1C).

Mutational signatures were extracted from the five children of the eBEACOPP-treated female. SBS clock-like signature was extracted from all the children, consistent with the previously reported mutational signatures in germline DNMs^23^. Additionally, the procarbazine-associated signature SBSB, was identified in all three children conceived post-chemotherapy and was absent in the two children born prior to chemotherapy.

Intriguingly, mutational signature analysis of sperm from the eBEACOPP-treated male revealed that approximately one third of mutations were attributed to SBSB (Figure 2B). The remaining mutations were mapped to SBS clock-like signatures as previously reported^18,24^.

To investigate whether ABVD, eBEACOPDac, or eBEACOPP exert consistent effects across multiple HSPC colonies, we examined the HSPC phylogenetic trees from all donors (Figure 3). The dacarbazine-associated signature, SBSC, was identified in all HSPC colonies of three ABVD-treated individuals and in 50-100% of colonies of four eBEACOPDac-treated individuals. In eBEACOPP-treated patients, the procarbazine-associated signatures, SBSA and SBSB, were jointly detected in all colonies, except in patient PD55816, where the signatures co-occurred on only two branches. HSPC colonies from normal donors only exhibit SBS clock-like signature (Extended Figure 2).

**Figure 3:**
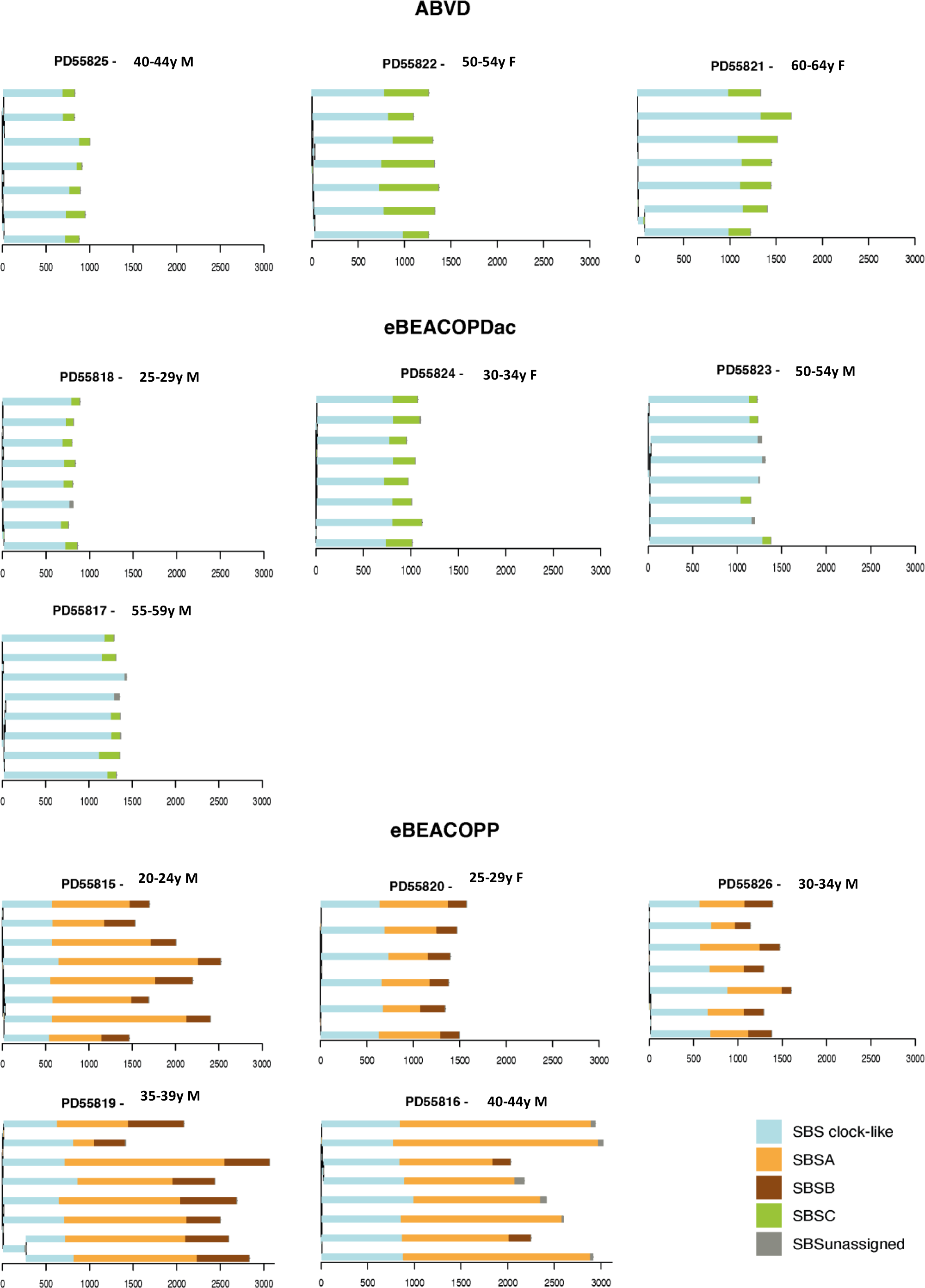
Phylogenetic trees with assigned mutational signatures from individuals treated with ABVD, eBEACOPDac or eBEACOPP. Individuals are ordered by age. Branch lengths are proportional to the mutation count. The mutational signatures contributing to each branch are colour coded as indicated at the bottom right. “SBSunassigned’’ indicates mutations that are not confidently attributed to any signature.

WGS of colon from a procarbazine-exposed patient revealed a significant burden of the procarbazine-associated signature, SBSA, in both normal colonic crypts and cancer micro-biopsies (Extended Figure 3). There is no increased frequency of SBSA in subclonal crypts relative to adjacent normal crypts, suggesting that mutations bearing this signature likely originated from a colorectal stem cell or occurred during early stages of tumorigenesis. These findings imply a long-term impact of procarbazine on normal solid tissues, beyond haematological effects.

The dacarbazine-associated signature, SBSC, was identified in all HSPC colonies of three ABVD-treated individuals. In BEACOPDac-treated patients, the presence of SBSC was observed in all colonies from patient PD55824, in 7/8 colonies from PD55818, in 6/8 colonies from PD55817, and in 4/8 colonies from PD55823. In eBEACOPP-treated patients, the procarbazine-associated signatures, SBSA and SBSB, were jointly detected in all colonies, except in patient PD55816, where the signatures co-occurred on only two branches.

### Clinical data analysis

From 2017 to 2022, 312 patients with advanced-stage cHL were treated with first-line eBEACOPDac at 25 centres in the UK, Ireland and France (Table S2). Their survival outcomes were compared with 1945 patients treated with eBEACOPP in the HD18 trial^20^. Using propensity score matching (PSM), 248 eBEACOPDac-treated HD18-like patients were matched with 248 patients treated with eBEACOPP in the HD18 trial. Patients were well-matched for age, sex, stage and IPS (Table S3). eBEACOPDac-treated patients received fewer cycles of chemotherapy than HD18 patients (median: 4 vs 6). 5.7% of the eBEACOPDac cohort received radiotherapy compared to 28.6% of the HD18 cohort. Median follow-up was 36 months (eBEACOPDac) and 57 months (HD18).

The 3-year PFS of the eBEACOPDac cohort mirrors the HD18 3-year PFS both before PSM (93.3% (CI_95_=90.3-96.4%) vs 93.3% (CI_95_=92.1-94.4%)) and after (92.1% (CI_95_=88.5-95.8%) vs 91.7% (CI_95_=88.1-95.3%)) (Figure 4). 3-year overall survival of the eBEACOPDac cohort is 99.3% (CI_95_=98.4-100%) (Supplementary Figure 1).

**Figure 4:**
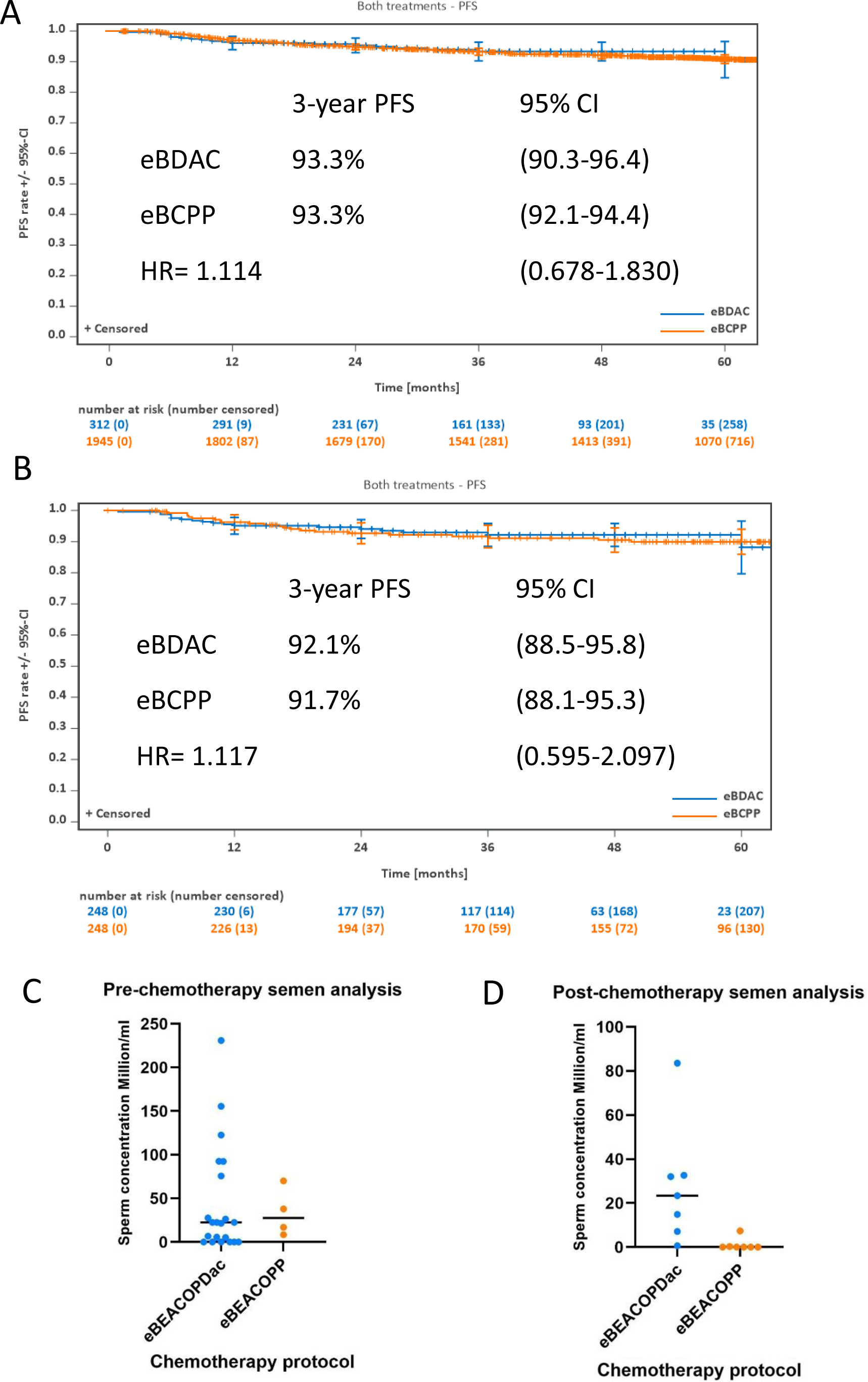
**(A)** Kaplan-Meier estimates of progression-free survival of the escalated BEACOPDac-treated patients (n=312) compared to the HD18 trial escalated BEACOPP-treated patients (n=1945) before propensity score matching. **(B)** Kaplan-Meier estimates of progression-free survival of the escalated BEACOPDac-treated patients (n=248) compared to the HD18 trial escalated BEACOPP-treated patients (n=248) after propensity score matching. The hazard ratio was obtained from Cox regression adjusted for propensity score. **(C)** Sperm concentration of Hodgkin lymphoma patients prior to treatment with escalated BEACOPDac or eBEACOPP chemotherapy. **(D)** Sperm concentration >2 years following completion of chemotherapy. eBPDac = escalated BEACOPDac, eBCPP = escalated BEACOPP, HR = Hazard ratio, PFS = progression-free survival, 95% CI = 95% Confidence interval

Of 312 patients who started eBEACOPDac, 3 patients had primary refractory disease, and 14 have relapsed at 6 to 60 months. 13 of 17 patients with relapse/refractory disease are currently in remission following subsequent treatment including haematopoietic stem cell transplantation (Supplementary Figure 2). One patient aged over 50 years died with bowel perforation during cycle 1 eBEACOPDac, one patient in his 30s died during allogeneic stem cell transplantation for relapsed disease, one patient aged over 30 years with alcoholic liver disease died in remission 8 months after eBEACOPDac, and one patient in his 40s died from suicide while in remission 3 years post-treatment for relapsed cHL.

From 2009 to 2022, 73 patients with advanced-stage cHL were treated with first-line eBEACOPP at 7 UK centres. Toxicity was compared between the eBEACOPDac cohort (n=312) and the real-world UK eBEACOPP cohort (n=73) over the first 4 cycles (Table S4). eBEACOPP and eBEACOPDac patients were well-matched with no significant differences in age, sex, or stage (stage 3/4 79% vs 83%), although the eBEACOPP patients had higher risk disease (IPS≥3 in 77% vs 62%; p=0.02). 57% of eBEACOPDac patients received only 4 cycles (vs 16% of eBEACOPP patients; p<0.001), as the eBEACOPDac cohort was largely treated after publication of HD18 trial data^20^. Most real-world eBEACOPP patients were treated with an HD15 approach^25^ (6 cycles of escalated BEACOPP followed by PET-guided radiotherapy).

When comparing the eBEACOPDac versus the eBEACOPP cohort, there was no significant difference in mean D8 ALT (46.0 vs 38.8; p=0.081); or in mean D8 neutrophil count (3.00 vs 2.55; p=0.125) in patients given granulocyte-colony stimulating factor from D9.

There were fewer non-elective days of inpatient care for eBEACOPDac patients compared to eBEACOPP patients (mean 3.32 vs 5.23; p=0.031). However, as eBEACOPDac patients were treated more recently there may be some era effect. Importantly, eBEACOPDac patients received fewer red cell transfusions compared to eBEACOPP patients (mean 1.72 vs 3.69; p<0.001). Of the women aged <35 years who completed ≥4 cycles of chemotherapy, 65/65 had a return of menstrual periods after eBEACOPDac compared to 25/28 after eBEACOPP. eBEACOPDac patients appeared to restart menstruation earlier post-chemotherapy (mean 5.04 vs 8.77 months; p=0.004). However, eBEACOPP patients received more cycles of chemotherapy. The use of Goserelin to suppress ovulation varied between centres.

eBEACOPDac and eBEACOPP-treated patients had similar sperm concentrations pre-treatment (median 22.5 vs 27.5 Million/ml, p=0.683; Figure 4C). However, >2 years post-chemotherapy, there was a striking difference between the two cohorts and 6/7 eBEACOPDac-treated males had a normal sperm concentration, while 6/7 eBEACOPP-treated patients were azoospermic (median 23.4 vs 0.0 Million/ml; p<0.01; Figure 4D).

## Conclusions

This is the first study demonstrating the impact of lymphoma polychemotherapy on genomic health of normal somatic tissues, tumour and the germline.

The SBS25 mutational signature has previously been described in two Hodgkin lymphoma cell lines from patients exposed to chemotherapy^26–28^, and has also been identified in cell-free DNA of treated cHL patients^29^. A recent study demonstrating SBS25 in relapsed cHL suggested a link with procarbazine/dacarbazine^30^. Our study has demonstrated that SBSA (very similar to SBS25), SBSB, and a novel indel signature are caused by procarbazine and contribute to the large excess mutation burden in the HSPCs of eBEACOPP-treated patients.

We have observed an increased number of de novo mutations and the procarbazine-associated signature, SBSB, in the sperm of an eBEACOPP-treated male, and in the germline DNA of children conceived after maternal procarbazine exposure. This observation marks the first direct detection of a chemotherapy signature in sperm, underscoring the potential long-term impact of treatment on germline DNA integrity. However, currently the duration of these effects in sperm and their potential implications for fertility are unknown. Our findings align with previous reports of chemotherapy-induced signatures in offspring^14^ and highlight the need for continued research into the hereditary implications of chemotherapy treatment.

The consequences of procarbazine-induced mutational burden and signatures in normal tissues remain unknown, however in some contexts procarbazine can cause the acquisition of driver mutations. Recent data from paediatric cHL survivors suggest that SBS25 is the likely cause of the *STAT3* Y640F mutation, which has previously been identified as a gain-of-function driver mutation in T-cell large granular lymphocytic leukaemia^31^. Our data indicate that SBS25 is not confined to HSPCs in procarbazine-exposed patients but is also induced in other stem cell compartments, including colonic mucosa^17^. However, as the driver mutations in this cHL survivor’s caecal adenocarcinoma were not in the T>A context, the role of SBS25 in this malignancy remains unclear.

Although not a prospective non-inferiority study, our data strongly suggest that replacing procarbazine with dacarbazine is unlikely to compromise the efficacy of eBEACOPP. There is no discernible difference in the 3-year PFS of the eBEACOPDac cohort and the HD18 trial cohort when comparing whole cohorts or in the sensitivity analysis. The data also indicate that replacing procarbazine with dacarbazine has toxicity benefits. Although the data are retrospective, they reveal a marked reduction in blood transfusion requirement in patients receiving eBEACOPDac compared to eBEACOPP (mean 1.72 vs 3.69 units; p<0.001), a finding that is mirrored closely in the prospective randomised HD21 trial, where patients treated with the procarbazine-free arm, BrECADD (brentuximab vedotin, etoposide, cyclophosphamide, doxorubicin, dacarbazine, dexamethasone), had a reduced blood transfusion requirement compared with eBEACOPP-treated patients (8% vs 22%)^32^.

This study also demonstrates a clear reduction in gonadal toxicity in patients treated with eBEACOPDac compared to those receiving eBEACOPP. Sperm concentration was normal >2 years post-chemotherapy in most eBEACOPDac-treated males, whereas there was a high incidence of azoospermia and oligospermia after eBEACOPP. These findings are consistent with previously published GHSG data^33^ and the EuroNet PHL C1 trial where the same drug substitution was made^4^. A return of menstrual periods was observed in all eBEACOPDac-treated females <35y and this occurred significantly earlier than in the real-world eBEACOPP cohort.

In conclusion, our study provides strong supportive evidence that eBEACOPDac is highly efficacious Hodgkin lymphoma therapy. The clear benefits in terms of stem cell genomic health and fertility demonstrated here provide even more reason for clinicians offering frontline eBEACOPP to consider replacing procarbazine with dacarbazine.

## Data Availability

All data produced in the present study are available upon reasonable request to the authors.

## Acknowledgements

This work was supported by the Addenbrooke’s Charitable Trust. Investigators at the Sanger Institute are supported by a core grant from the Wellcome Trust. Work in the EL laboratory is supported by a core grant from the Wellcome Trust and Medical Research Council to the Cambridge Stem Cell Institute and by a Wellcome Trust – Royal Society Sir Henry Dale Fellowship. We thank the investigators at the Sanger Institute for the whole genome sequencing of HSPCs, sperm and buccal DNA for this study. We are grateful to the donors for tissue donation.

This research was funded in whole, or in part, by the Wellcome Trust. For the purpose of open access, the author has applied a Creative Commons Attribution (CC BY) licence to any Author Accepted Manuscript version arising from this submission.

## Data

The sample identifiers contained in this manuscript are not known to anyone outside of the research group. Age ranges are included to avoid direct identifiers. Please contact the corresponding author if precise ages or further information required.

