## Supplementary Appendix for "Procarbazine-induced Genomic Toxicity in Hodgkin Lymphoma Survivors"

**Supplementary Methods**

**Extended Figure 1**

**Extended Figure 2**

**Extended Figure 3**

**Supplementary Figure 1**

**Supplementary Figure 2**

**Supplementary Figure 3**

**Table S1**

**Table S2**

**Table S3**

**Table S4**

**References**

### Supplementary Methods:

#### Cohort and sample preparation

Patients donated blood for research to the Cambridge Blood and Stem Cell Biobank (CBSB), which has Health Research Authority and NHS Research Ethics Committee approval (REC reference 18/EE/0199, IRAS 149581).

Peripheral blood mononuclear cells (PBMC) were isolated from 12 advanced-stage Hodgkin lymphoma patients who had been in remission for ≥6 months. The patients had been previously treated with either eBEACOPDac (6 cycles (n=2); 4 cycles (n=2)), eBEACOPP (6 cycles (n=3); 5 cycles (n=1); 4 cycles (n=1)) or ABVD (n=3; all 6 cycles). PBMCs were cultured for 14 days and single-cell derived HSPC colonies were harvested. DNA was extracted from each colony and 6-8 single-cell derived HSPC colonies were whole-genome sequenced from each of the 12 patients (n=91; mean sequencing depth 26X). CaVEMan[1], used for calling single nucleotide variants (SNVs), was run against an unmatched synthetic normal genome. Single HSPC somatic mutation burdens and signatures from the chemotherapy-exposed individuals were compared to those from the previously published normal cohort (n=110; mean sequencing depth 24X)[2].

Insertions and deletions (indels) were identified using the Pindel algorithm applied to a matched normal sample. The same dataset-specific filters previously described for substitutions[3] were also utilised for indel analysis. Next, indels underwent genotyping with following criteria: a variant allele frequency (VAF) exceeding 0.2, a minimum sequencing depth of 10, and at least five mutant reads.

Whole-genome sequencing was also performed on a caecal adenocarcinoma from a Hodgkin lymphoma survivor treated with Chlorambucil, Vinblastine, Procarbazine, Prednisolone

(ChlVPP) nine years before sampling[4]. Previously published[4] normal colorectal epithelium from the same individual was also interrogated.

The linear regression of age and SNV or indel mutation burden[2] from the control cohort was used as a baseline against which to compare the mutation burden of the chemotherapy-exposed individuals.

**Multi-sibling family study of a eBEACOPP-treated cHL female**

Given it is not feasible to directly study the effect of chemotherapy treatment on oocytes in female patients, buccal DNA was obtained from five children and the spouse of a cHL female patient who was treated with eBEACOPP prior to conceiving the 3rd-5th children. Buccal DNA was extracted using the Qiagen DNA investigator kit and subjected to whole-genome sequencing (with median coverage ranging from 28X to 34X). SNVs were called using CaVEMan[1], running against an unmatched synthetic normal. The artefacts were removed by filtering out variants with 1) if the median alignment score of reads supporting a mutation less than 140; 2) If more than half of the reads aligned to the position were clipped; 3) a cruciform filter was also applied to remove artefacts introduced during amplification of cruciform DNA. Germline DNMs were called by genotyping the variants across the parents and the children and applying further filters by removing 1) sites that had more than 10% of reads supporting the alternative allele in either of the parents; 2) sites with read depth in the top 0.01% quantile. assuming read depth to be Poisson distributed, with the $\lambda$ parameter of the Poisson distribution equal to the mean genome read depth; 3) sites with less than two reads supporting the alternative allele on each strand; 4) SNVs with less than 30% variant allele fraction. Mutational burden and signatures were compared between pre- (n=2) and post-chemotherapy children (n=3). Using the 2-mutation variance between the two eldest children (both pre-chemotherapy) and assuming an 80% paternal origin for these mutations[5], the annual increase in paternally acquired DNMs was estimated at 1.6. Subsequently, we calculated the expected paternal mutations for the post-chemotherapy children using the following equation.

$$Ni = Ni\text{-}1 + (1.6 * (Ai - Ai\text{-}1))$$

Where N represents the number of paternal de novo mutations; A signifies the age of the father at conception, and i denotes the child number (ordered by descending age). We further calculated the total expected de novo mutations by dividing the above counts by 0.8. The expected de novo mutations and the observed de novo mutations in the post-chemotherapy children were compared using t-test.

We further phased the de novo mutations from the offsprings to confirm the increase in the maternally inherited mutations. Germline mutations for each trio were jointly identified using GATK HaplotypeCaller v4.2.4.1[6], and the de novo mutations were phased using PhaseMyDeNovo (https://github.com/queenjobo/PhaseMyDeNovo).

**Sequencing of the sperm sample from eBEACOPP patient**

The sperm DNA (200 ng) of a patient with mild oligospermia (13M/ml) after undergoing 4 cycles of eBEACOPP treatment was sequenced using Nanoseq whole-genome sequencing[7], achieving a mean duplex coverage of 7.5 dx. The sequencing data underwent analysis using the NanoSeq variant calling pipeline (https://github.com/cancerit/NanoSeq). SNVs (n=194) were identified using established filters[7]. The mutation burden was corrected by dividing the number of mutations called by the total duplex coverage of the sample. To establish a control cohort for comparison, the parental germline de novo point mutation burden was predicted based on pedigree studies from parents and offspring trio whole-genome sequencing[5].

**Mutational signature analysis**

The Hierarchical Dirichlet process (HDP), a method based on Bayesian hierarchical Dirichlet processes (available at https://github.com/nicolaroberts/hdp), was employed to extract mutational signatures in this study. HDP was utilised without prior assumptions on single base substitutions (SBS) /Indels, derived from phylogenetic trees obtained from HSPCs, de novo germline mutations from the offspring of a female who received 6 cycles of eBEACOPP, and mutations from the sperm sample of a eBEACOPP-treated male in his 30s. Since the sperm data was generated using Nanoseq method, corrections for trinucleotide context abundance were applied to the NanoSeq mutations. The mutations were adjusted by multiplying with the ratio of the duplex coverage distribution for the 32 pyrimidine trinucleotide context to the genome-wide distribution.

The HDP analysis was performed with dual hierarchies: chemotherapy treatment (with "normal" used for untreated individuals) and individual id. Both clustering hyperparameters, alpha and beta, were set to one. The Gibbs sampler was executed with 30,000 iterations, a spacing of 200 iterations, and 100 collected iterations. After each iteration, three iterations of concentration parameters were performed. Six components were initially extracted, with two components excluded as noise.

When compared to the COSMIC v3.4 signatures, one of the HDP components represented a blend of previously documented clock-like signatures. Other three components were identified as de novo signatures predominantly present in individuals who underwent chemotherapy. Only signatures contributing more than 5% of the mutation burden in the sample were considered. Signatures with less than 5% contribution were classified as overfitting and their contribution was categorised under "Unassigned."

Indel signatures were also extracted from indels identified in HSPCs using HDP with the same aforementioned parameters. Two components were extracted, of which one represented a mixture of previously known clock-like signatures. The second component was specifically associated with individuals who had undergone eBEACOPP treatment. Once again, only signatures contributing more than 5% were deemed active.

**Clinical data analysis**

This is a retrospective study of 312 patients with advanced-stage cHL, treated with first-line escalated BEACOPDac at 25 centres in the UK, Ireland and France between 2017 and 2022. eBEACOPDac is a modified version of the eBEACOPP protocol, in which oral procarbazine is removed and replaced with intravenous dacarbazine (250mg/m$^2$ D2-3) (Table S1).

Progression-free survival (PFS) of the 312 eBEACOPDac-treated patients were compared with 1945 eBEACOPP-treated HD18 trial patients[8], through collaboration with the German Hodgkin Study Group (GHSG). The majority (265/312) of eBEACOPDac patients were treated using an 'HD18-like' approach[8], namely those with a metabolic remission on interim PET-CT after 2 cycles (iPET2) received 2 additional cycles of eBEACOPDac, and those with positive iPET2 received 4 additional cycles of eBEACOPDac. To refine our comparative analysis, we used propensity score matching (PSM) based on age, sex, stage, and international prognostic score (IPS) to match HD18-like patients with a subgroup of HD18 trial patients. PFS was calculated from the date of diagnosis to the date of progression, death, or last follow-up. Survival analyses were performed using the Kaplan-Meier method.

Toxicity outcomes of the eBEACOPDac cohort were compared with a real-world eBEACOPP cohort of 73 patients treated at 7 UK centres between 2009 and 2022. Toxicity data were collected from patients who had received ≥4 cycles of chemotherapy. The outcomes studied were day 8 (D8) neutrophil count, D8 alanine transaminase (ALT), units of red cells transfused, and days of non-elective hospital admission. In women aged <35 years who had received ≥4 cycles of chemotherapy, data were collected on use of the gonadotrophin-releasing hormone (GnRH) agonist Goserelin, and on time from completion of chemotherapy to return of menstrual periods. Sperm concentration data were collected pre-treatment and ≥2 years post-chemotherapy.

Statistical analyses were performed using R software and SPSS. The Mann-Whitney U test and t-test were used for continuous variables and Fisher's exact test for categorical variables. The study was conducted with Health Research Authority and Public Benefit and Privacy Panel approval.

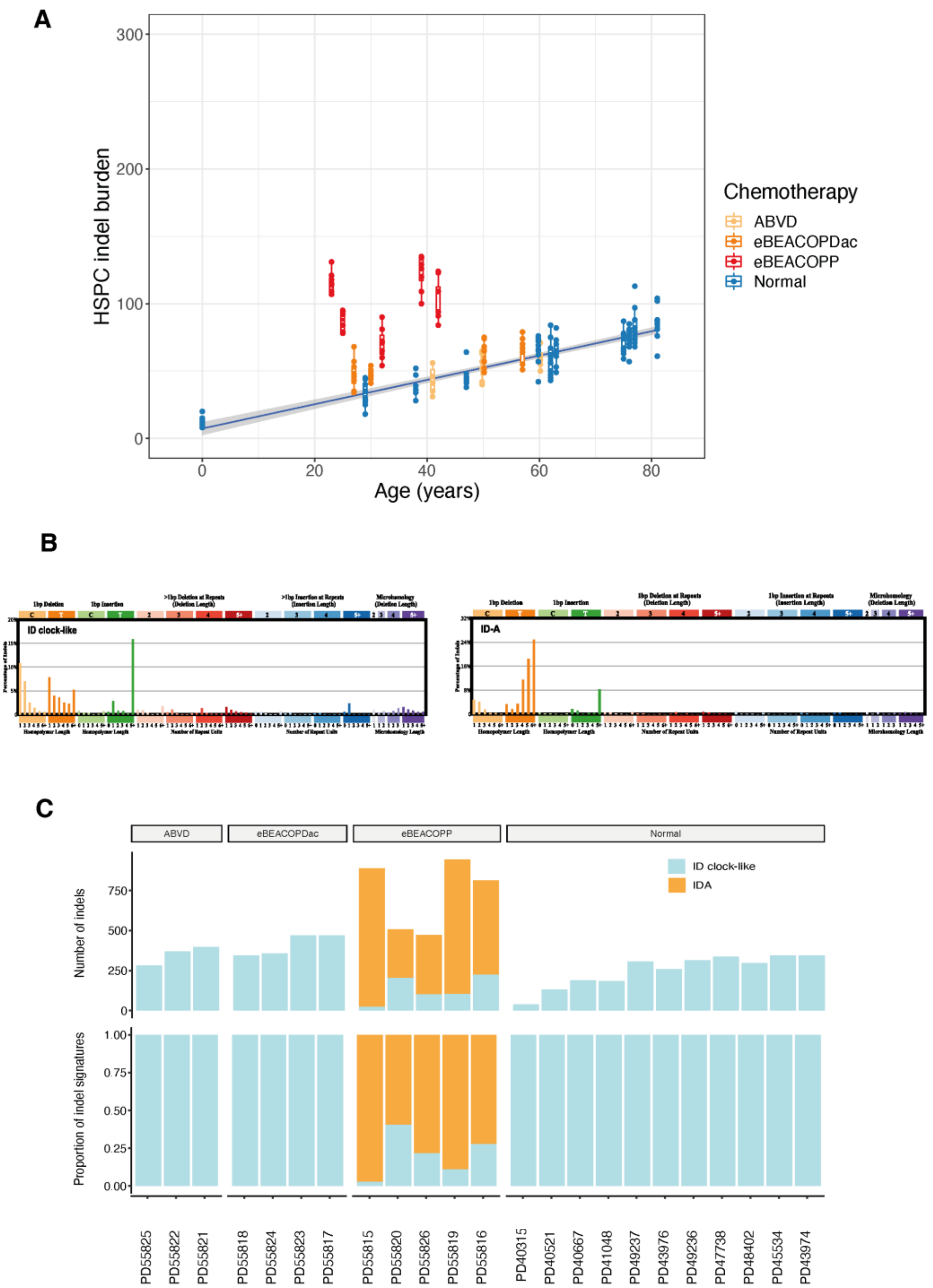


**<u>Extended Figure 1:</u> (A)** Burden of HSPC indels across the chemotherapy donor and a comparable normal donor cohort. The boxplots represent data from individual HSPC colonies (n = 201; 6-10 colonies per individual) and are coloured by chemotherapy exposure. The boxes indicate the median and interquartile range and the whiskers denote the range. The blue line represents a regression of age on mutation burden for the normal donors, with 95% CI shaded in grey. **(B)** Mutational signatures extracted using HDP. **(C)** Proportion of extracted signatures active in eBEACOPP, cBEACOPDac, ABVD and normal individuals.

HSPC=Haematopoietic stem and progenitor cells, indels=insertions and deletions, HDP=Hierarchical Dirichlet process

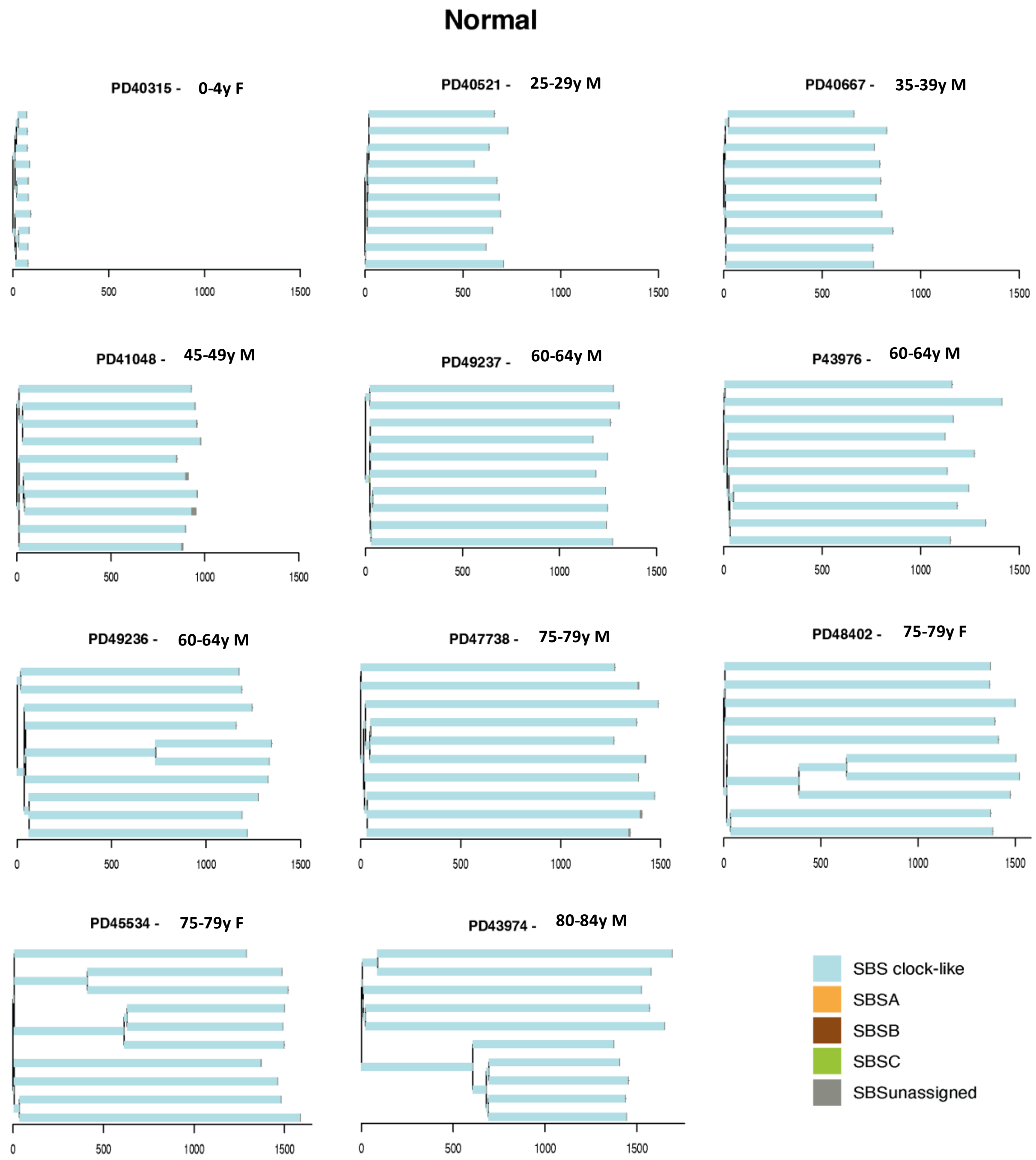


**Extended Figure 2:** Phylogenetic trees with assigned mutational signatures from 11 normal donors. Individuals are ordered by age. Branch lengths are proportional to the mutation count. The mutational signatures contributing to each branch are colour coded as indicated at the bottom right. "SBSunassigned " indicates mutations that are not confidently attributed to any signature.

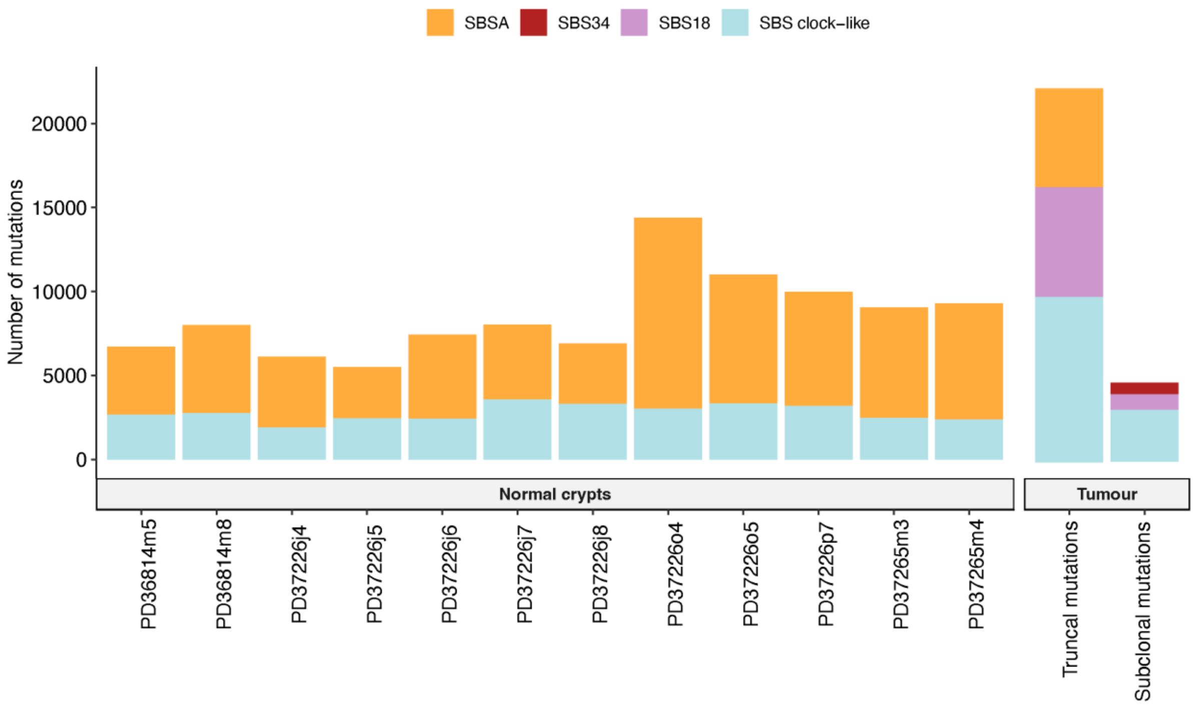


**<u>Extended Figure 3:</u>** The stacked bar plot illustrates the number of mutations contributing to different mutational signatures in colonic crypts from a Hodgkin lymphoma patient with caecal adenocarcinoma diagnosed 9 years after treatment with ChIVPP/PABIOE. Different colours reflect the four signatures extracted (SBS clock-like, SBS18, SBS34 and SBSA). There are 12 normal crypts, compared with tumour truncal mutations and tumour subclonal mutations. The truncal mutations are found in every single tumour cell, while the subclonal mutations are present only in a fraction of the tumour cells.

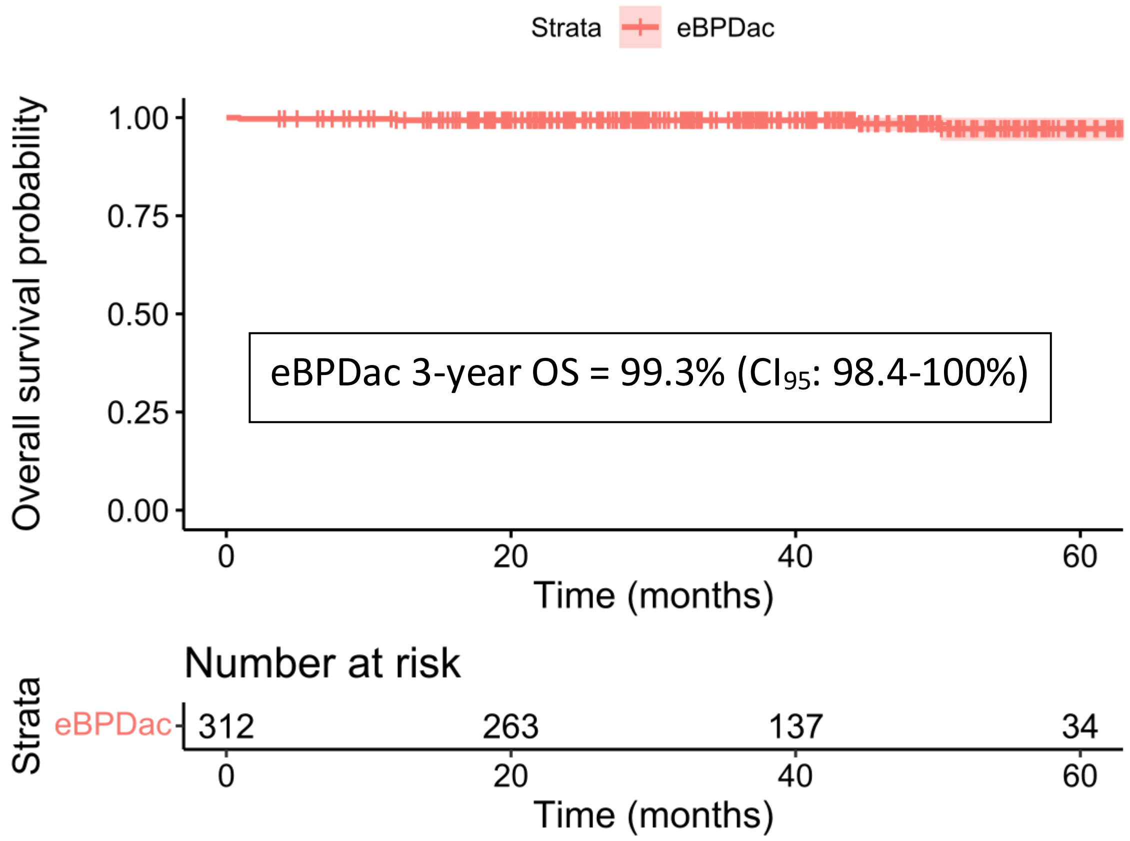


**<u>Supplementary Figure 1:</u>** Kaplan-Meier estimate of overall survival of the escalated BEACOPDac-treated patients (n=312). Red shading represents 95% confidence interval.

eBPDac=escalated BEACOPDac, OS=overall survival.

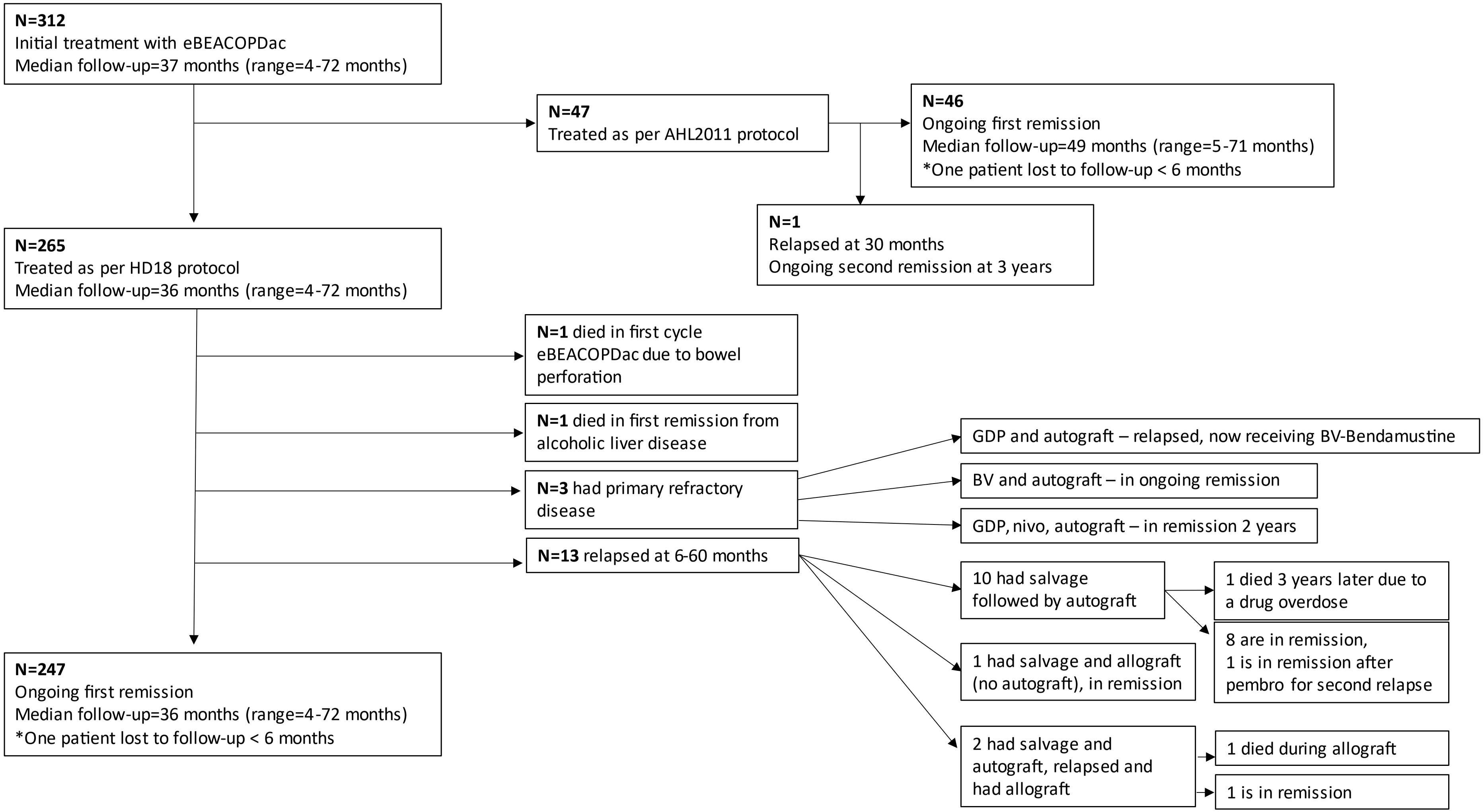


**<u>Supplementary Figure 2:</u> Consort diagram of outcomes of 312 patients treated with first line eBEACOPDac for advanced stage Hodgkin lymphoma.**

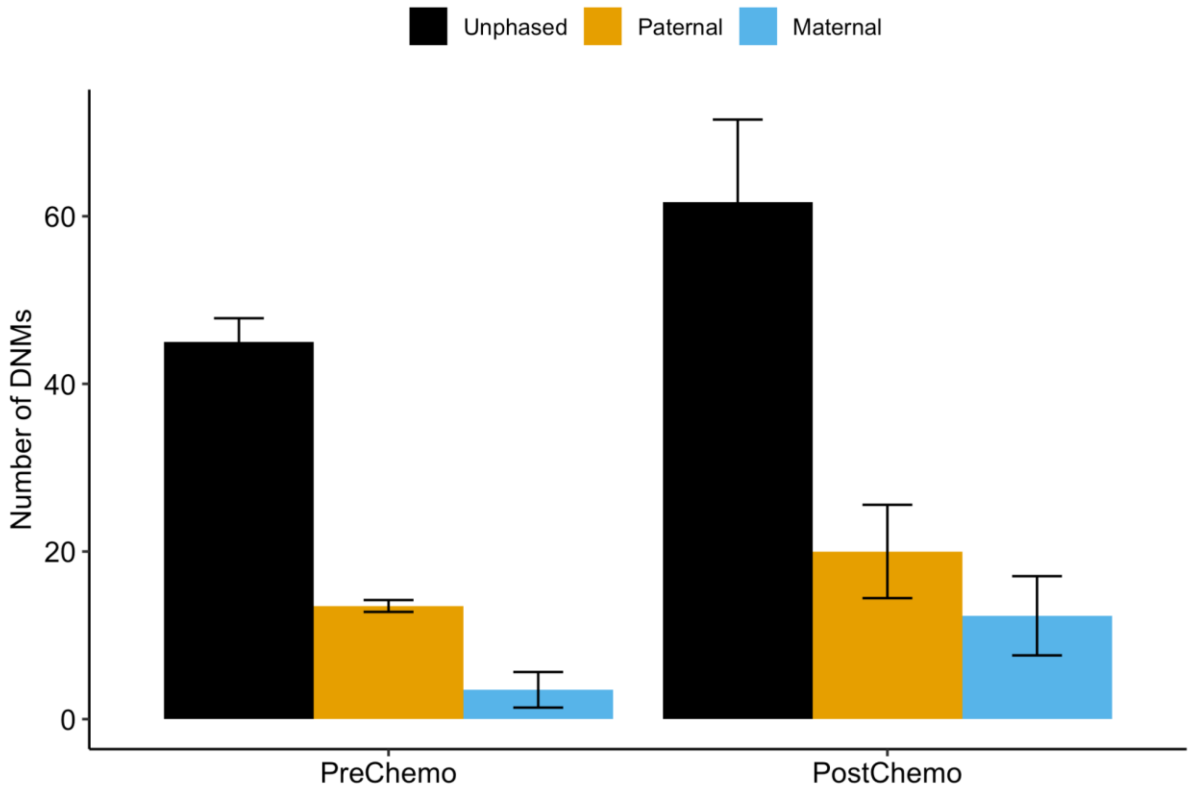


**Supplementary Figure 3:** Read-based phasing of DNMs shows an excess (~2.3 fold) of maternally-derived DNMs in post-chemotherapy children of the eBEACOPP-treated cHL female.

DNMs=De novo mutations, cH=Classical Hodgkin lymphoma

**<u>Table S1:</u> Escalated BEACOPDac treatment protocol**

| | | | | |
|---|---|---|---|---|
| Cyclophosphamide | 1250 mg/m$^2$ | i.v. | Day 1 | Over 60 min |
| Doxorubicin | 35 mg/m$^2$ | i.v. | Day 1 | Over 30 min |
| Etoposide | 200 mg/m$^2$ | i.v. | Day 1-3 | Over 60 min |
| Dacarbazine | 250 mg/m$^2$ | i.v. | Day 2-3 | Over 30 min |
| Prednisolone | 40 mg/m$^2$ | p.o. | Day 1-14 | |
| Vincristine | 1.4 mg/m$^2$ (max 2mg) | i.v. | Day 8 | i.v. bolus |
| Bleomycin | 10 mg/m$^2$ | i.v. | Day 8 | i.v. bolus |
| GCSF (Filgrastim) | 300mcg | s.c. | Day 9-12 | |

Repeat on day 22.

**<u>Table S2:</u> Baseline characteristics (before propensity score matching) of escalated BEACOPDac patients (n=312) versus HD18 trial escalated BEACOPP patients (n=1945). eBEACOPDac patients also shown subdivided into HD18-like (n=265) and AHL2011 (n=47) approaches to treatment. HD18 trial patients suitable for PSM (n=973) are shown, this sample is based on patients recruited after trial amendment and with at ≥1 cycle of treatment in the full-ITT collective.**

| **Baseline Characteristics** | **Escalated BEACOPDac** **N=312** | **eBEACOPDac patients** treated as per HD18 protocol **n=265** | **eBEACOPDac patients** treated as per AHL2011 protocol **n=47** | **HD18 trial patients** **N=1945** | **HD18 trial patients** Suitable for PSM **N=973** |
|---|---|---|---|---|---|
| Median Age, years (range) | 26 (16-62) | 25 (16-62) | 29 (20-59) | 32 (18-60) | 32 (18-60) |
| Age≤ 45 years | 288 (92.3%) | 246 (92.8%) | 42 (89.4%) | 1563 (80.4%) | 765 (78.6%) |
| Age >45 years | 24 (7.7%) | 19 (7.2%) | 5 (10.6%) | 382 (19.6%) | 208 (21.4%) |
| Male sex (%) | 186 (59.6%) | 162 (61.1%) | 24 (51.1%) | 1183 (60.8%) | 589 (60.5%) |
| Stage 2B / 2X / 2XB | 52 (16.6%) | 41 (15.5%) | 11 (23.4%) | Stage I-III 1237 (63.6%) | Stage I-III 604 (62.1%) |
| 3 | 50 (16.0%) | 45 (16.9%) | 5 (10.6%) | | |
| 4 | 210 (67.3%) | 179 (67.5%) | 31 (66.0%) | 708 (36.4%) | 369 (37.9%) |
| IPS 0-2 | 117 (37.6%) | 93 (35.2%) | 24 (51.1%) | 1207 (62.3%) | 604 (62.1%) |
| 3-4 | 152 (48.8%) | 132 (50.0%) | 20 (42.5%) | | |
| 5-7 | 42 (13.5%) | 39 (14.8%) | 3 (6.4%) | | |
| IPS ≥3 | 194 (62.4%) | 171 (64.8%) | 23 (48.9%) | 731 (37.7%) | 368 (37.8%) |
| Unknown | 1 | 1 | | | |
| Total cycle number | | | | | |
| 1 | 4 (1.3%) | 4 (1.5%) | 0 (0.0%) | | |
| 2 | 46 (14.7%) | 1 (0.4%) | 45 (95.7%) | | |
| 4 | 178 (57.1%) | 178 (67.2%) | 0 (0.0%) | | |
| 5 | 5 (1.6%) | 5 (1.9%) | 0 (0.0%) | | |
| 6 | 79 (25.3%) | 77 (29.1%) | 2 (4.2%) | | |
| Median cycle number | 4 (1-6) | 4 (1-6) | 2 (2-6) | 6 (2-8) | 6 (2-8) |
| iPET Deauville score | | | | | |
| 1- 2 | 89 (28.9%) | 75 (28.7%) | 14 (29.7%) | 1005 (51.7%) | 437 (46.2%) |
| 3 | 145 (47.1%) | 121 (46.3%) | 24 (51.1%) | 471 (24.2%) | 275 (29.0%) |
| 4 | 72 (23.4%) | 63 (24.1%) | 9 (19.1%) | 469 (24.1%) | 235 (24.8%) |
| 5 | 2 (0.6%) | 2 (0.8%) | 0 (0.0%) | | |
| ≤3 | 234 (76.0%) | 196 (75.1%) | 38 (80.9%) | 1476 (75.9%) | 712 (75.2%) |
| No iPET | 4 | 4 | | | 26 |
| Radiotherapy | 16 (5.2%) | 14 (6.0%) | 2 (4.3%) | 385 (19.2%) | 218 (22.0%) |
| Progressive disease | 18 (5.8%) | 17 (6.4%) | 1 (2.1%) | 128 (6.6%) | 65 (6.7%) |
| Death | 4 (1.3%) | 4 (1.5%) | 0 (0.0%) | 79 (4.1%) | 32 (3.3%) |
| Follow-up, months Median (min-max) | 36.9 (0.9-72.7) | 35.9 (0.9-72.7) | 49.1 (5.0-71.1) | 65.5 (1.5-133.7) | 58.2 (0.4-96.7) |

**<u>Table S3:</u> Baseline characteristics (after propensity score matching) of escalated BEACOPDac patients (n=248) versus HD18 trial escalated BEACOPP patients (n=248)**

| Patient Characteristics | Escalated BEACOPDac (after PSM) N=248 | | HD18 Escalated BEACOPP (after PSM) N=248 | |
|---|---|---|---|---|
| Median Age, years (range) | 25.5 (16-62) | | 25 (18-58) | |
| Age ≤ 45 years<br>Age >45 years | 229 (92.3%)<br>19 (7.7%) | | 225 (90.7%)<br>23 (9.3%) | |
| Male sex (%) | 157 (63.3%) | | 157 (63.3%) | |
| Stage I-III<br>IV | 85 (34.3%)<br>163 (65.7%) | | 91 (36.7%)<br>157 (63.3%) | |
| IPS 0-2<br>IPS >2 | 93 (37.5%)<br>155 (62.5%) | | 91 (36.7%)<br>157 (63.3%) | |
| Median number of cycles, (range) | 4 (1-6) | | 6 (1-8) | |
| Number of cycles | PET pos | PET neg | PET pos | PET neg |
| 1-3 | 2 (3.3%) | 2 (1.0%) | 3 (2.1%) | 1 (1.0%) |
| 4 | 0 (0.0%) | 167 (90.3%) | 0 (0.0%) | 50 (50.5%) |
| 5 | 3 (5.0%) | 2 (1.0%) | 1 (0.7%) | 1 (1.0%) |
| 6 | 55 (91.7%) | 14 (7.6%) | 133 (93.7%) | 47 (47.5%) |
| 7-8 | 0 (0.0%) | 0 (0.0%) | 5 (3.5%) | 0 (0.0%) |
| N | 60 | 185 | 142 | 99 |
| Radiotherapy | 14/246 (5.7%) | | 71 (28.6%) | |
| Progressive disease/relapse | 16 (5.8%) | | 21 (8.5%) | |
| Death | 4 (1.6%) | | 3 (1.2%) | |
| Median follow-up in months (range) | 36.0 (0.9-72.7) | | 57.3 (1.5-95.2) | |

PSM = Propensity score matching, IPS = International Prognostic Score.

**Table S4: Baseline characteristics and treatment outcomes of real-world UK escalated BEACOPP cohort (n=73) and escalated BEACOPDac cohort (n=312)**

| **Baseline Characteristics** | **eBEACOPP N=73** | **eBEACOPDac N=312** | **p-value** |
|---|---|---|---|
| Median Age, years (range)<br><br>Male sex (%) | 26 (16-57)<br><br>37 (50.7%) | 26 (16-62)<br><br>186 (59.6%) | U=11228, p=0.852<br><br>Fisher, p=0.188 |
| Stage 2B / 2X / 2XB<br>3<br>4 | 15 (20.5%)<br>9 (12.3%)<br>49 (67.1%) | 52 (16.7%)<br>50 (16.0%)<br>210 (67.3%) | (Stage 2-3 vs 4)<br>Fisher, p=1 |
| IPS 0-2<br>3-4<br>5-7<br>IPS ≥3<br>Unknown | 17 (23.3%)<br>40 (54.7%)<br>16 (21.9%)<br>56 (76.7%)<br>0 | 117 (37.6%)<br>152 (48.9%)<br>42 (13.5%)<br>194 (62.3%)<br>1 | (IPS 0-2 vs ≥3)<br>Fisher, **p=0.0208** |
| **Treatment Outcomes** | | | |
| Total cycle number 1<br>2<br>4<br>5<br>6<br>7<br>8 | 1 (1.4%)<br>0 (0.0%)<br>12 (16.4%)<br>5 (6.8%)<br>53 (72.6%)<br>1 (1.4%)<br>1 (1.4%) | 4 (1.2%)<br>46 (14.7%)<br>178 (57.1%)<br>5 (1.6%)<br>79 (25.3%)<br>0 (0.0%)<br>0 (0.0%) | Fisher,<br>(4 vs 6) **p=9.92E-14** |
| iPET Deauville score 1- 2<br>3<br>4<br>5<br>≤3<br>No iPET | 16 (25.8%)<br>27 (43.5%)<br>19 (30.6%)<br>0 (0.0%)<br>43 (69%)<br>11 | 89 (28.8%)<br>145 (47.1%)<br>72 (23.3%)<br>2 (0.6%)<br>234 (76.0%)<br>4 | (DS ≤3 vs 4-5)<br>Fisher,<br>p=0.267 |
| Mean day 8 ALT U/L (cycles 1-4) [SD]<br><br>Mean day 8 neutrophils × $10^9$/L (cycles 1-4) [SD]<br>*(GCSF day 9, eBPP n=66, eBPDac n=169)*<br><br>*(GCSF day 4, eBPP n=1, eBPDac n=87)* | 38.8 (±29.5)<br><br><br>3.00 (±1.95)<br><br>6.93 (NA) | 46.0 (±29.2)<br><br><br>2.55 (±2.18)<br><br>6.29 (±6.22) | t(100)=1.76<br>p=0.081<br><br>t(131)=1.54, p=0.125 |
| Mean days non-elective admission (cycles 1-4) [SD]<br><br>Mean number of red cell units transfused (cycles 1-4) [SD] | 5.23 (±7.23)<br><br><br>3.69 (±3.89) | 3.23 (±5.96)<br><br><br>1.70 (±2.77) | U=7688,<br>**p=0.031**<br><br>U=5815<br>**p=7.40E-7** |
| Median follow-up, months (range) | 72.1 (5.0-153) | 30.0 (0.92-71.1) | U=3518, **p=2.2E-16** |
| Mean number of months for return of menstrual period post-chemotherapy [SD]<br>(eBPDac N=65, eBPP N=28)<br>Mean number of cycles completed by women <35 years of age [SD] | 8.77 (±5.57)<br><br><br>5.74(±0.59) | 5.04 (±3.07)<br><br><br>4.63 (±0.93) | t(30)=3.16, **p=0.00357**<br><br>U=459, **p=2.92E-7** |
| Median sperm concentration Million/ml:<br>Pre-treatment (range) *eBPP n=4, eBPDac n=21*<br>>2 years post-chemotherapy (range)<br>*eBPP n=7, eBPDac n=7* | <br>27.5 (8.5-70.0)<br><br>0.0 (0.0-7.4) | <br>22.5 (0.0-231.0)<br><br>23.4 (0.6-83.5) | <br>U=36, p=0.683<br><br>U=47, **p=0.00404** |
